# Psychiatric Characteristics and Mental Health Service Use Among Hospitalised Youth in Foster Care, Adoption, and Child Welfare Supervision: A Retrospective Comparative Study

**DOI:** 10.64898/2026.09.13.26362960

**Authors:** Mireia Solerdelcoll, Gisela Sugranyes, Inmaculada Baeza

**Author notes:** **Corresponding author:** Mireia Solerdelcoll and Gisela Sugranyes,Child and Adolescent Psychiatry and Psychology Department, Institute of Neurosciences, Hospital Clínic de Barcelona, 170 Villarroel, 08036 Barcelona, Spain, (M. Solerdelcoll); (G. Sugranyes). Both authors contributed equally as co-corresponding authors (Mireia Solerdelcoll, Gisela Sugranyes).

## Abstract

**Background:** Youth involved in child protection systems are at increased risk of psychiatric disorders and are overrepresented in inpatient mental health services. However, few studies have directly compared the clinical characteristics and mental health service use of youth across different child protection pathways.

**Objective:** To compare clinical characteristics, psychosocial risk factors, clinical complexity, and mental health service use among psychiatrically hospitalised youth in foster care, adoption, and child welfare supervision.

**Participants and Setting:** This retrospective observational study included 236 children and adolescents across three child protection pathways admitted to a Child and Adolescent Inpatient Psychiatry Unit between 2018 and 2024.

**Methods:** Clinical, psychosocial, and service-use data were extracted from medical records and compared using univariate analyses, multilevel mixed-effects models, logistic regression, and time-to-event analyses.

**Results:** All groups showed high comorbidity and clinical severity, but distinct clinical profiles emerged. Youth in foster care more frequently presented with externalising disorders, trauma exposure, and substance use; adopted youth showed higher rates of neurodevelopmental and prenatal adversity-related conditions; and youth under child welfare supervision more often presented with affective symptoms and suicidal ideation. Post-discharge emergency department re-consultation differed significantly between groups (*p* = 0.007) and was most frequent among adopted youth, who also had higher odds of psychiatric readmission. Youth in foster care had the shortest median time to emergency re-consultation, although time-to-event differences were not significant (*p* = 0.476).

**Conclusions:** Distinct clinical and service-use profiles across child protection pathways support trauma-informed, placement-sensitive mental health care and stronger coordination between child protection and mental health services.

## Introduction

Children and adolescents involved in child protection systems (CPS)—including foster care (FC), adoption, and child welfare supervision—represent one of the most vulnerable populations in society. These youth are at particularly high risk of psychopathology, owing not only to early-life adversities such as perinatal complications, neglect, abuse, institutional rearing, and family instability (Bronsard et al., 2016; Humphreys et al., 2015), but also to underlying genetic vulnerability (Nielsen et al., 2025). Offspring of parents with psychiatric disorders have more than twice the risk of experiencing maltreatment, highlighting the complex interplay between inherited and environmental influences (Assink et al., 2018; Baldwin et al., 2023). Consequently, CPS-involved youth are disproportionately represented in specialist mental health services, including inpatient psychiatric care (Benarous et al., 2023; Chorniy et al., 2026; Kronström et al., 2023).

Child welfare supervision aims primarily to preserve family unity through supportive monitoring, preventive interventions, and oversight of care, whereas FC provides temporary out-of-home placements when a child’s safety cannot be ensured at home. Such placements are often characterised by instability, disrupted attachments, and limited continuity in care (Tarren-Sweeney, 2008). Adoption, in contrast, offers a permanent family environment but frequently follows early institutional deprivation, prenatal adversity, and disruptions in caregiving during sensitive developmental periods— particularly among children adopted at older ages or through international adoption. Despite shared developmental risks, clinical needs and outcomes may diverge depending on exposure to adversity, age at intervention, placement stability, and access to post-placement support (van IJzendoorn et al., 2020).

Extensive research has shown that youth involved in CPS exhibit a substantially higher burden of mental health difficulties compared with their peers in the general population, with markedly elevated rates of internalising and externalising disorders, substance use disorders, trauma-related symptoms, and neurodevelopmental conditions (Bronsard et al., 2016; Humphreys et al., 2015; Seker et al., 2022).

Youth in FC frequently present with complex, comorbid clinical profiles, shaped by cumulative adversity, repeated disruptions in caregiving, and placement instability (Ford et al., 2007; Oswald et al., 2010; Tarren-Sweeney, 2008). Adopted youth are also vulnerable to persistent emotional and behavioural difficulties, even when raised in stable post-adoption environments (Juffer & van IJzendoorn, 2005). These difficulties are particularly pronounced among those adopted later in childhood or following early institutional care (Palacios et al., 2019; Kumsta et al., 2010). Neurodevelopmental conditions such as autism spectrum disorder (ASD) and foetal alcohol spectrum disorders (FASD) show elevated prevalence among internationally adopted children—particularly those from Eastern Europe and Russia—and among children with histories of institutionalisation (Landgren et al., 2010; Palacios & Brodzinsky, 2010; Rodriguez-Perez et al., 2023). Youth under child welfare supervision—who remain with their families under social service monitoring—have received limited empirical attention; however, available evidence suggests substantial rates of internalising and externalising difficulties (Burns et al., 2004; Jud et al., 2016).

Despite these findings, few studies have directly compared the psychiatric profiles and service needs of youth across the three principal CPS pathways. Most prior research has focused on single groups or limited pairwise comparisons, constraining understanding of shared and distinct developmental trajectories (Lehmann, 2021; Solerdelcoll et al., 2026; Tarren-Sweeney & Hazell, 2006). Moreover, most evidence derives from community or population samples, with limited attention to youth requiring specialist inpatient psychiatric care (Fontanella et al., 2020). In a previous study, we examined the psychiatric characteristics and service use of youth in FC admitted to an inpatient psychiatric unit compared with a non-foster-care control group (Solerdelcoll et al., 2022). The current study extends this work by incorporating adopted youth and those under child welfare supervision at the time of psychiatric hospitalisation, enabling a broader and more nuanced comparative analysis across the three main CPS pathways.

### Study Aims

This study aimed to compare clinical characteristics and mental health service use among youth admitted to inpatient psychiatric care under three child protection arrangements: FC, adoption, and child welfare supervision. Specifically, we examined psychosocial risk factors, psychiatric diagnoses, and patterns of mental health service use across groups. Based on previous research, we hypothesised that youth in FC would present with higher rates of externalising and substance use disorders, greater exposure to adversity, and greater clinical complexity than youth in the other groups. We further hypothesised that adopted youth would show a greater prevalence of neurodevelopmental disorders and conditions associated with prenatal adversity. Given the limited comparative evidence regarding youth under child welfare supervision and service-use outcomes, analyses involving these domains were considered exploratory.

## Methods

### Study design and sample

This retrospective observational study included all patients aged under 18 years admitted to the Child and Adolescent Inpatient Psychiatry Unit of Hospital Clínic de Barcelona between January 2018 and December 2024 who were under one of the following child protection arrangements at the time of admission: (1) institutional FC; (2) adoption; or (3) child welfare supervision, referring to youth who remained with their families under the supervision of the regional child protection teams in Catalonia, Spain (*Equips d’Atenció a la Infància i l’Adolescència*; Direcció General d’Atenció a la Infància i l’Adolescència, n.d.). For individuals with multiple admissions during the study period, only the first hospital admission was included. Youth with temporary protection measures or unclear child protection status were excluded. The study was approved by the Clinical Research Ethics Committee of Hospital Clínic de Barcelona. Given the retrospective, non-interventional design of the study, no direct or indirect risks to participants were anticipated.

### Procedures and assessments

Electronic medical records were reviewed systematically by a child and adolescent psychiatrist. Relevant data were extracted using a structured data abstraction form, anonymised, and entered into an electronic database.

Sociodemographic and clinical variables were systematically recorded, including age, sex assigned at birth, country of birth, and living arrangements. Clinical records were reviewed for documented history of maltreatment (physical, emotional, or sexual abuse, or neglect), peer victimisation, perinatal adversity, and pharmacological treatment at discharge.

Information on family history psychiatric disorders, substance use disorders, and intellectual disability was collected from clinical records.

For youth in FC, placement trajectory data were extracted, including total number of placements, age at first placement, previous family foster care, and status as unaccompanied migrant minors. For adopted youth, variables included age at adoption, international versus domestic adoption, country of origin, pre-adoptive background, and post-adoption disruptions. When available, these data were supplemented with information from caregivers, referring professionals, and primary mental health care providers.

The primary reason for admission was recorded based on the clinician’s judgement at the time of initial assessment. Psychiatric diagnoses at discharge were assigned by the treating consultant child and adolescent psychiatrist, according to *Diagnostic and Statistical Manual of Mental Disorders, Fifth Edition* (DSM-5) criteria. Participants meeting criteria for psychosis risk syndrome, based on ultra-high-risk criteria supplemented by attenuated negative symptoms and extended genetic-risk criteria, were identified and recorded as a separate diagnosis. The assessment procedures and inclusion criteria have been described previously (Dolz et al., 2019).

Service-related variables included referral source, discharge destination, number of psychiatric admissions, length of hospital stay, and frequency of emergency department use. Post-discharge mental health service use was assessed using the time elapsed between discharge and the first emergency department visit, and the time to psychiatric readmission, measured in days, as indicators of clinical recurrence and relapse risk.

## Statistical analysis

Statistical analyses were conducted using SPSS Statistics version 28.0 (IBM Corp.). Descriptive statistics summarised socio-demographic, clinical, and service-related variables. Continuous variables were reported as means with standard deviations or as medians with interquartile ranges (IQR), depending on their distribution. Categorical variables were presented as frequencies and percentages.

Group differences were examined using chi-square (χ²) tests for categorical variables, with Fisher’s exact test applied where expected cell counts were <5. Effect sizes were estimated using Cramer’s V. For continuous variables, one-way analysis of variance (ANOVA) or Kruskal–Wallis tests were applied depending on normality assumptions. Post hoc comparisons were performed using Mann–Whitney U tests. Effect sizes were reported as *r* (correlation coefficient), indicating the magnitude of differences (small ≥ 0.1, medium ≥ 0.3, large ≥ 0.5).

Patterns of emergency department use and psychiatric readmission before and after hospitalisation were analysed using multilevel mixed-effects linear regression models, with group, time (pre/post), and group-by-time interaction as fixed effects, and patient ID as a random effect to account for within-subject variation. Time-to-event outcomes (first emergency department visit and psychiatric readmission after discharge) were analysed with Cox proportional hazards models. Group differences in readmission risk were estimated using logistic regression. Given the age-related distribution of substance use and psychiatric disorders, secondary analyses restricted to youth aged over 12 years were conducted for diagnostic and clinical complexity outcomes.

Clinical complexity was assessed using two indicators: psychiatric comorbidity, defined as the number of discharge diagnoses and dichotomised as ≥2 versus <2, and polypharmacy, defined as the prescription of ≥2 psychotropic classes at discharge. Group comparisons were conducted using χ² tests for categorical variables and ANOVA or Kruskal–Wallis tests for continuous variables. All tests were two-tailed, with statistical significance set at *p* < 0.05.

## Results

### 1. Sample characteristics

A total of 236 patients were included: foster care (FC; n = 96), adoption (n = 60), and child welfare supervision (n = 80). Sex distribution did not differ significantly between groups (*p* = 0.479), with a female predominance across all groups (FC: 59.4%; adoption: 55.0%; child welfare supervision: 65.0%). Age at first psychiatric hospitalisation differed significantly (*p* = 0.006, η² = 0.036), with adoptees admitted at an older mean age (15.86 ± 1.52 years) than those under FC (14.89 ± 2.17 years) or child welfare supervision (15.01 ± 1.86 years). Country of birth varied significantly between groups (*p* < 0.001): most FC (49.0%) and child welfare supervision (60.0%) youth were Spanish-born, whereas adoptees were primarily born in Russia (31.7%) or in other Eastern European countries (11.7%). A substantial proportion of FC youth were born in Morocco (22.9%).

Family history of psychiatric disorders did not differ significantly between groups (*p* = 0.091), although information on biological family psychiatric history was unavailable for 58.3% of adopted youth. In contrast, significant group differences emerged for family history of substance use disorders (*p* < 0.001), which were most frequently reported among adoptees with available family history information, compared with youth in FC and under child welfare supervision.

History of interpersonal victimisation (maltreatment and/or bullying) was highly prevalent across the sample and differed significantly between groups *(p* < 0.001), with the highest rates observed among youth under child welfare supervision (78.8%), followed by FC youth (70.8%), while adopted youth showed a substantially lower rate (45.0%) (see Table 1).

**Table 1.** Sociodemographic and child protection characteristics (N = 236)

| Variable | FC<br>(n = 96) | Adoption<br>(n = 60) | CWS<br>(n = 80) | Statistic | p-value | Effect size |
| --- | --- | --- | --- | --- | --- | --- |
| Age, mean (SD)<sup>a</sup> | 14.89 ± 2.17<br>[8–17] | 15.86 ± 1.52<br>[11–17] | 15.01 ± 1.86<br>[9–17] | F(2,233) =<br>5.21 | 0.006* | η<sup>2</sup> = 0.036 |
| Female sex, n (%) | 57 (59.4%) | 33 (55.0%) | 52 (65.0%) | χ<sup>2</sup>(2) = 1.47 | 0.479 | V = 0.08 |
| Country of birth, n (%)<sup>b</sup> |  |  |  | χ<sup>2</sup>(14) = 106.82 | < 0.001* | V = .476 |
| Spanish-born, n (%) | 47 (49.0%) | 11 (18.3%) | 48 (60.0%) |  |  |  |
| Born in Russia or Eastern Europe, n (%) | 3 (3.1%) | 26 (43.3%) | 1 (1.3%) |  |  |  |
| Born in Morocco, n (%) | 22 (22.9%) | 4 (6.7%) | 3 (3.8%) |  |  |  |
| Parents separated, n (%) | 50 (52.1%) | 12 (20.0%) | 60 (75.0%) | χ<sup>2</sup>(2) = 44.35 | < 0.001* | V = 0.434 |
| Family psychiatric history, n (%) |  |  |  |  |  |  |
| Any psychiatric disorder<sup>c</sup> | 42 (58.3%) | 9 (36.0%) | 47 (60.3%) | χ<sup>2</sup>(2) = 5.99 | 0.091 | V = 0.17 |
| Substance use disorders | 34 (47.2%) | 22 (88.0%) | 34 (43.6%) | χ<sup>2</sup>(2) = 20.43 | < 0.001* | V = 0.30 |
| Unknown | 24 (25.0%) | 35 (58.3%) | 2 (2.5%) | χ<sup>2</sup>(2) = 97.73 | < 0.001* | V = 0.49 |
| History of interpersonal victimisation<sup>d</sup>, n (%) | 68 (70.8%) | 27 (45.0%) | 63 (78.8%) | χ<sup>2</sup>(2) = 28.41 | < 0.001* | V = 0.31 |
| Unaccompanied migrant minor, n (%) | 13 (13.5%) | 0 | 0 | χ<sup>2</sup>(2) = 20.06 | < 0.001* | V = 0.292 |

Note. FC = foster care; CWS = child welfare supervision; SD = standard deviation.
<sup>a</sup> Age at first psychiatric admission
<sup>b</sup> The omnibus chi-square test included all eight country-of-birth categories.
<sup>c</sup> Comprises any psychiatric disorder in a first- or second-degree relative, including mood, psychotic, anxiety and behavioural disorders.
<sup>d</sup> History of interpersonal victimisation refers to documented exposure to severe maltreatment (physical, sexual or emotional abuse and/or neglect) and/or peer victimisation (bullying).

Child protection trajectories revealed that FC youth entered care at a mean age of 12.9 years (SD = 3.0); 24% were placed in FC for the first time following hospital discharge, one-third had previously experienced family foster care (mainly kinship placements), and 13.5% were unaccompanied migrant minors. Family breakdown was documented as the reason for placement in 20.8% of cases. Among adopted youth, 68.3% had experienced institutional care prior to adoption, with a mean age at adoption of 35.3 months (SD = 27.2). Post-adoption, 86.7% remained with their adoptive family, though a minority experienced subsequent placement disruptions. Adopted youth showed significantly higher rates of prematurity and perinatal complications compared with the other groups. In the child welfare supervision group, most youth remained with their biological families under protective supervision, although 16.3% experienced residential care during the study period. Supervision was most commonly initiated due to neglect, parental conflict, or parental psychiatric/substance use problems (see Table 2).

**Table 2A.**
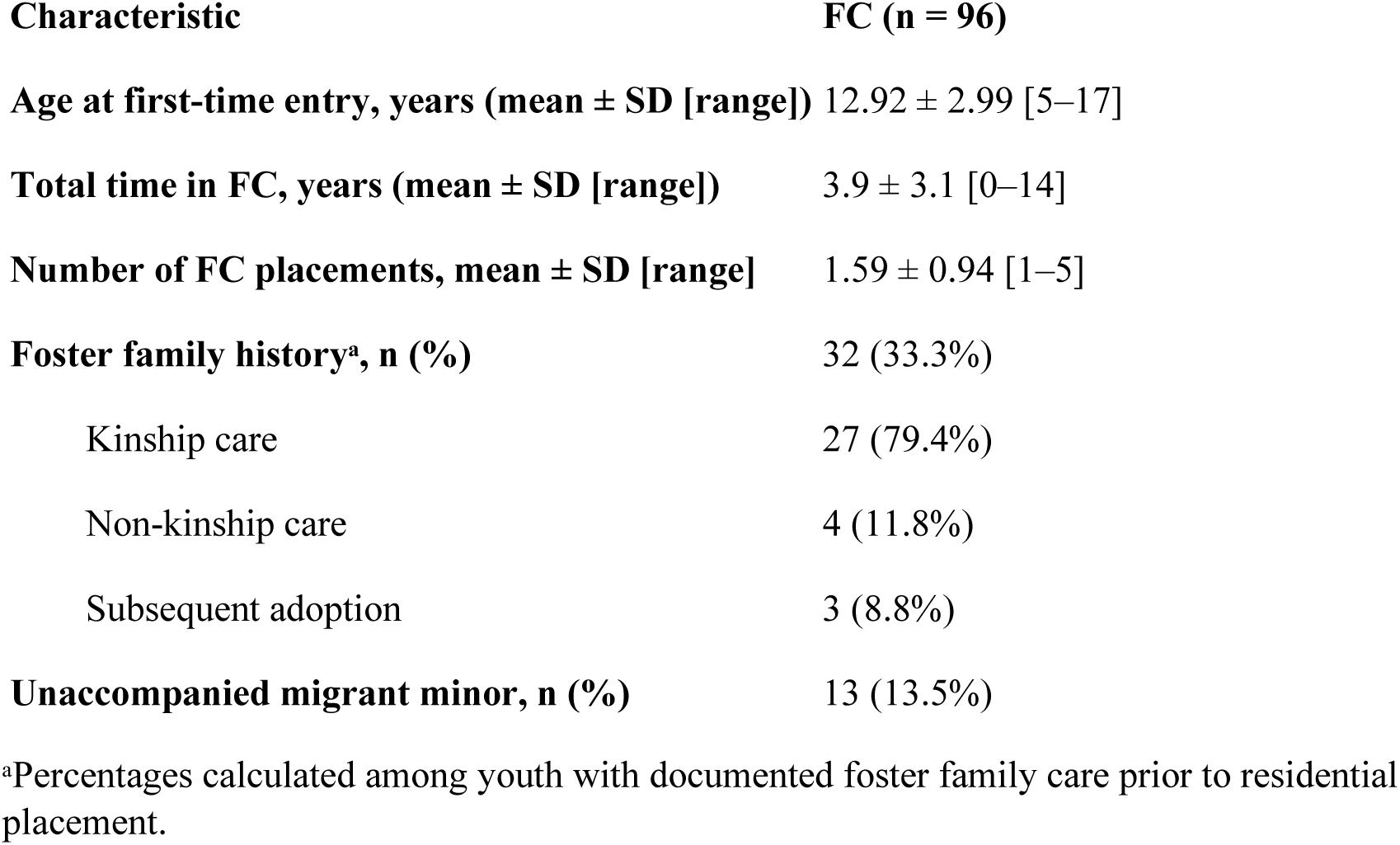
Placement characteristics among youth in Foster Care (FC)

| **Characteristic** | **FC (n = 96)** |
| --- | --- |
| **Age at first-time entry, years (mean ± SD [range])** | 12.92 ± 2.99 [5–17] |
| **Total time in FC, years (mean ± SD [range])** | 3.9 ± 3.1 [0–14] |
| **Number of FC placements, mean ± SD [range]** | 1.59 ± 0.94 [1–5] |
| **Foster family history<sup>a</sup>, n (%)** | 32 (33.3%) |
| Kinship care | 27 (79.4%) |
| Non-kinship care | 4 (11.8%) |
| Subsequent adoption | 3 (8.8%) |
| **Unaccompanied migrant minor, n (%)** | 13 (13.5%) |

<sup>a</sup>Percentages calculated among youth with documented foster family care prior to residential placement.

**Table 2B.** Adoption-related characteristics among adopted youth Characteristic Adoption (n = 60) Age at adoption, months (mean ± SD [range]) 35.27 ± 27.24 [0–120] Pre-adoption care settings, n (%)

| **Characteristic** | **Adoption (n = 60)** |
| --- | --- |
| **Age at adoption, months (mean ± SD [range])** | 35.27 ± 27.24 [0–120] |
| **Pre-adoption care settings, n (%)** |  |
| Biological family | 9 (15.0%) |
| Kinship care | 1 (1.7%) |
| Non-kinship care | 4 (6.7%) |
| Foster care | 5 (8.3%) |
| Institutional care (orphanage) | 41 (68.3%) |
| **Post-adoption continuity, n (%)** |  |
| Remain with adoptive family | 52 (86.7%) |
| Non-kinship care | 1 (1.7%) |
| Foster care | 4 (6.7%) |
| Homelessness | 3 (5.0%) |
| **Perinatal complications, n (%)<sup>\*</sup>** | 17 (28.3%) |
| **Nutritional deficits, n (%)** | 6 (10.0%) |

<sup>\*</sup>Including prematurity.

**Table 2C.** Placement characteristics among youth under child welfare supervision (CWS) Characteristic CWS (n = 80)

| **Characteristic** | **CWS (n = 80)** |
| --- | --- |
| **Kinship care involvement during CWS, n (%)** | 5 (6.3%) |
| **Any placement in FC, n (%)** | 11 (13.8%) |
| Before admission * | 5 (6.3%) |
| After admission | 6 (7.5%) |

\* Foster care placement occurred outside the index hospitalisation period (either before or after admission).

### 2. Clinical features

Primary reasons for psychiatric admission did not differ significantly between groups (*p* = 0.574). Behavioural and emotional dysregulation was the most frequent reason for admission among youth in FC (41.7%) and adopted youth (35.0%), whereas suicidal ideation or attempts were the most frequent reason among youth under child welfare supervision (35.0%) (Table 3A).

**Table 3.**
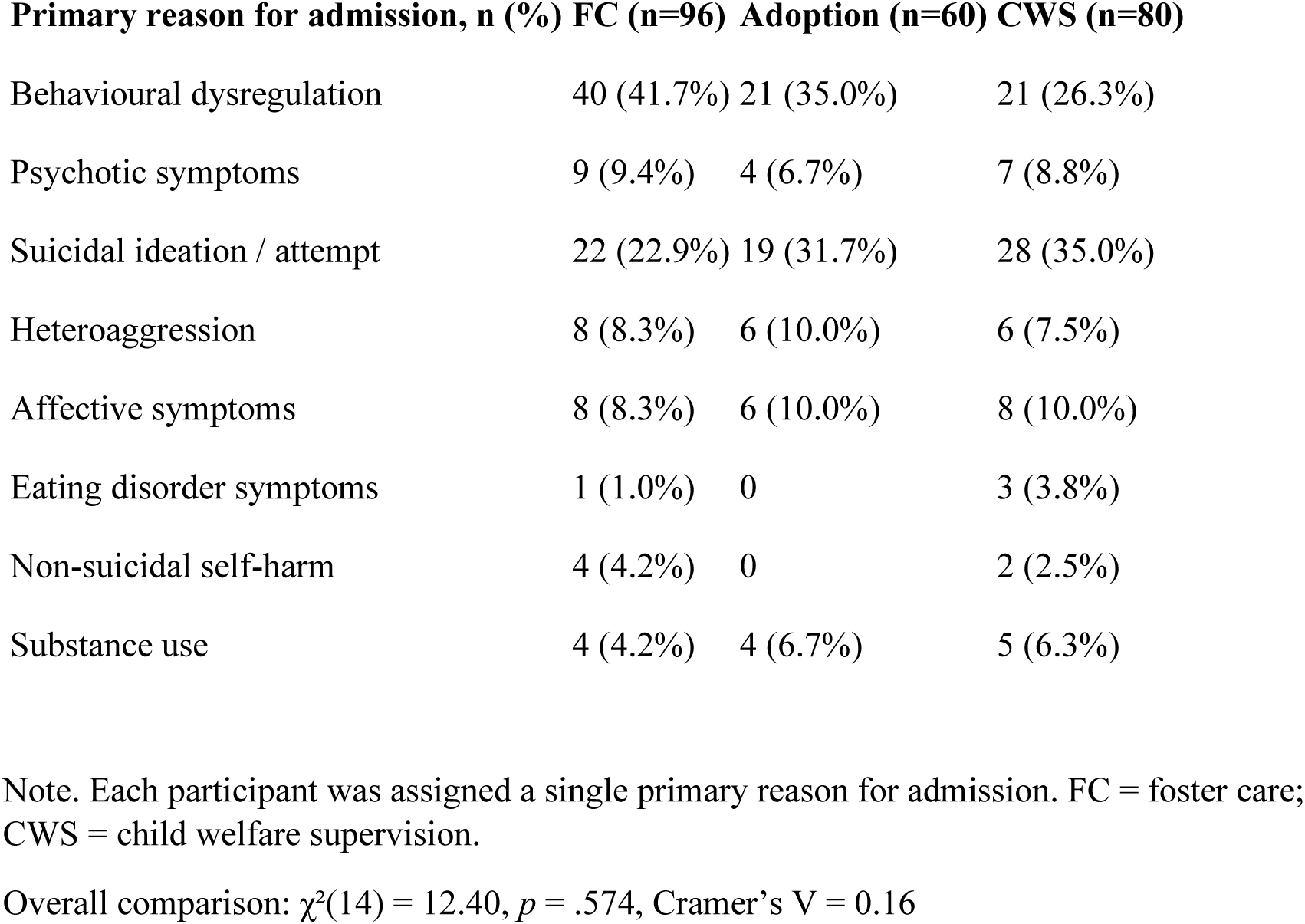
Reasons for psychiatric admission, DSM-5 diagnoses at discharge, and pharmacological treatment. **Table 3A. Primary reasons for current psychiatric admission**

**Table 3B.**
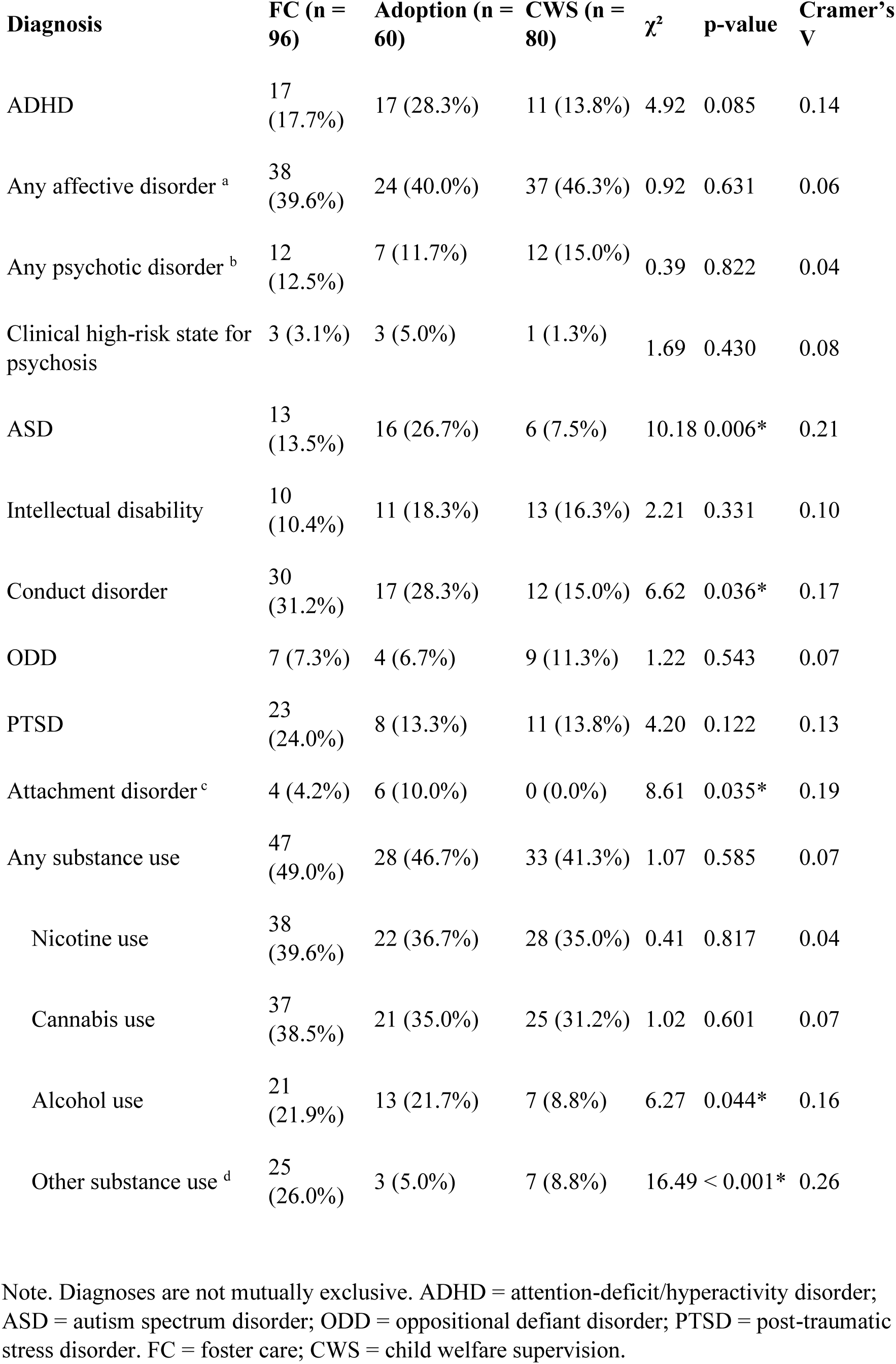
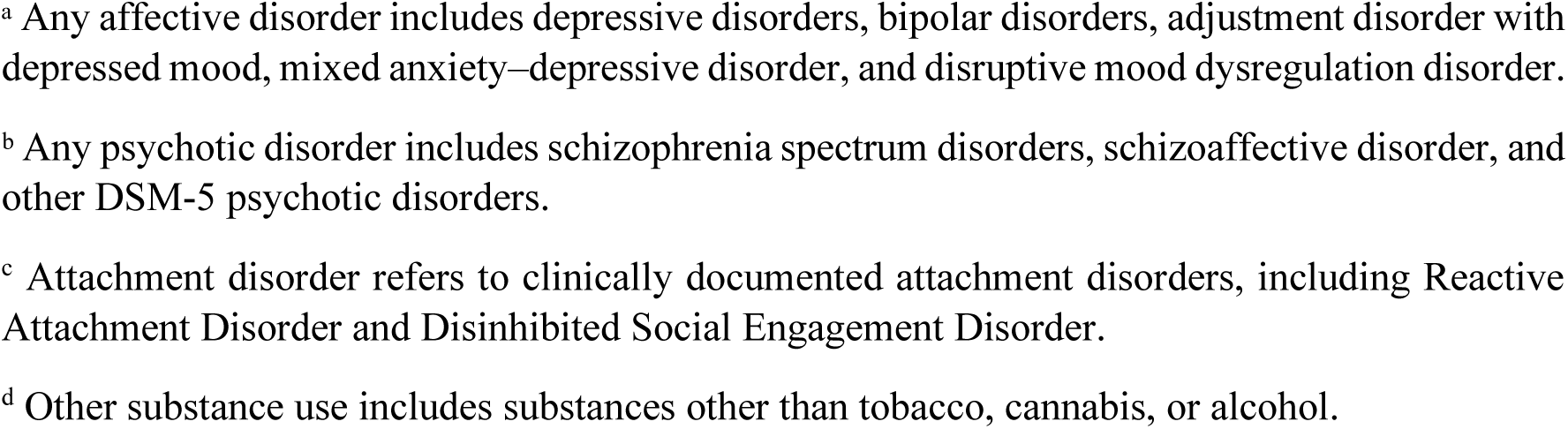
Psychiatric diagnoses at discharge.

| **Diagnosis** | **FC (n = 96)** | **Adoption (n = 60)** | **CWS (n = 80)** | **$\chi^2$** | **p-value** | **Cramer's V** |
| --- | --- | --- | --- | --- | --- | --- |
| ADHD | 17 (17.7%) | 17 (28.3%) | 11 (13.8%) | 4.92 | 0.085 | 0.14 |
| Any affective disorder <sup>a</sup> | 38 (39.6%) | 24 (40.0%) | 37 (46.3%) | 0.92 | 0.631 | 0.06 |
| Any psychotic disorder <sup>b</sup> | 12 (12.5%) | 7 (11.7%) | 12 (15.0%) | 0.39 | 0.822 | 0.04 |
| Clinical high-risk state for psychosis | 3 (3.1%) | 3 (5.0%) | 1 (1.3%) | 1.69 | 0.430 | 0.08 |
| ASD | 13 (13.5%) | 16 (26.7%) | 6 (7.5%) | 10.18 | 0.006* | 0.21 |
| Intellectual disability | 10 (10.4%) | 11 (18.3%) | 13 (16.3%) | 2.21 | 0.331 | 0.10 |
| Conduct disorder | 30 (31.2%) | 17 (28.3%) | 12 (15.0%) | 6.62 | 0.036* | 0.17 |
| ODD | 7 (7.3%) | 4 (6.7%) | 9 (11.3%) | 1.22 | 0.543 | 0.07 |
| PTSD | 23 (24.0%) | 8 (13.3%) | 11 (13.8%) | 4.20 | 0.122 | 0.13 |
| Attachment disorder <sup>c</sup> | 4 (4.2%) | 6 (10.0%) | 0 (0.0%) | 8.61 | 0.035* | 0.19 |
| Any substance use | 47 (49.0%) | 28 (46.7%) | 33 (41.3%) | 1.07 | 0.585 | 0.07 |
| Nicotine use | 38 (39.6%) | 22 (36.7%) | 28 (35.0%) | 0.41 | 0.817 | 0.04 |
| Cannabis use | 37 (38.5%) | 21 (35.0%) | 25 (31.2%) | 1.02 | 0.601 | 0.07 |
| Alcohol use | 21 (21.9%) | 13 (21.7%) | 7 (8.8%) | 6.27 | 0.044* | 0.16 |
| Other substance use <sup>d</sup> | 25 (26.0%) | 3 (5.0%) | 7 (8.8%) | 16.49 | < 0.001* | 0.26 |

Note. Diagnoses are not mutually exclusive. ADHD = attention-deficit/hyperactivity disorder; ASD = autism spectrum disorder; ODD = oppositional defiant disorder; PTSD = post-traumatic stress disorder. FC = foster care; CWS = child welfare supervision.
<sup>a</sup> Any affective disorder includes depressive disorders, bipolar disorders, adjustment disorder with depressed mood, mixed anxiety–depressive disorder, and disruptive mood dysregulation disorder.
<sup>b</sup> Any psychotic disorder includes schizophrenia spectrum disorders, schizoaffective disorder, and other DSM-5 psychotic disorders.
<sup>c</sup> Attachment disorder refers to clinically documented attachment disorders, including Reactive Attachment Disorder and Disinhibited Social Engagement Disorder.
<sup>d</sup> Other substance use includes substances other than tobacco, cannabis, or alcohol.

**Table 3C.**
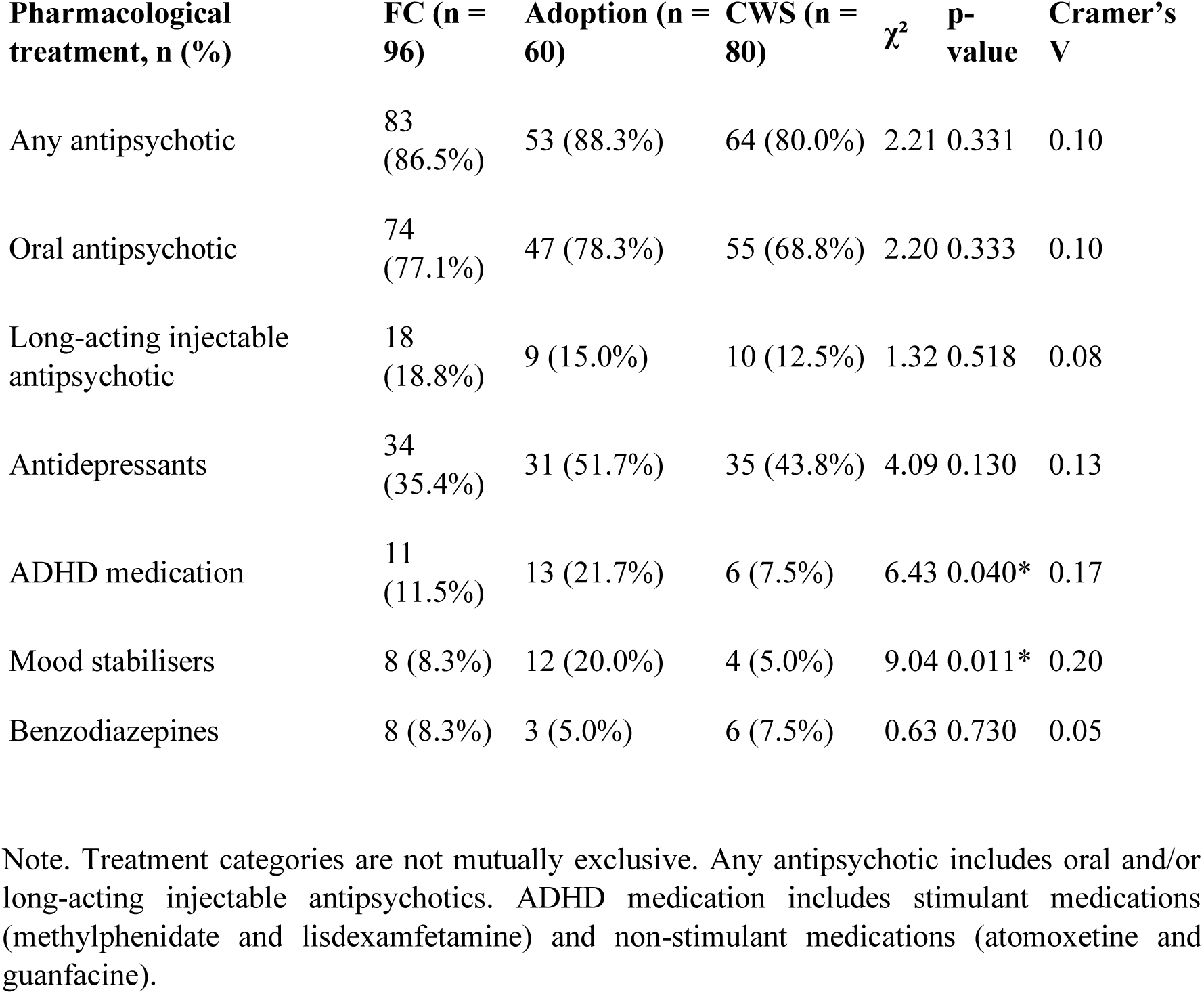
Pharmacological treatment at discharge.

| Pharmacological treatment, n (%) | FC (n = 96) | Adoption (n = 60) | CWS (n = 80) | $\chi^2$ | p-value | Cramer's V |
| --- | --- | --- | --- | --- | --- | --- |
| Any antipsychotic | 83 (86.5%) | 53 (88.3%) | 64 (80.0%) | 2.21 | 0.331 | 0.10 |
| Oral antipsychotic | 74 (77.1%) | 47 (78.3%) | 55 (68.8%) | 2.20 | 0.333 | 0.10 |
| Long-acting injectable antipsychotic | 18 (18.8%) | 9 (15.0%) | 10 (12.5%) | 1.32 | 0.518 | 0.08 |
| Antidepressants | 34 (35.4%) | 31 (51.7%) | 35 (43.8%) | 4.09 | 0.130 | 0.13 |
| ADHD medication | 11 (11.5%) | 13 (21.7%) | 6 (7.5%) | 6.43 | 0.040* | 0.17 |
| Mood stabilisers | 8 (8.3%) | 12 (20.0%) | 4 (5.0%) | 9.04 | 0.011* | 0.20 |
| Benzodiazepines | 8 (8.3%) | 3 (5.0%) | 6 (7.5%) | 0.63 | 0.730 | 0.05 |

Note. Treatment categories are not mutually exclusive. Any antipsychotic includes oral and/or long-acting injectable antipsychotics. ADHD medication includes stimulant medications (methylphenidate and lisdexamfetamine) and non-stimulant medications (atomoxetine and guanfacine).

Across all groups, the most frequent diagnoses were attention-deficit/hyperactivity disorder (ADHD), mood and psychotic disorders, disruptive behaviour disorders, and substance use disorders. Significant between-group differences emerged: ASD (*p* = 0.006) and attachment disorder (*p* = 0.035) were more frequent among adoptees, whereas conduct disorder (*p* = 0.036), alcohol use (*p* = 0.044), and other substance use (*p* < 0.001) were most frequent among FC youth (see Table 3B). Secondary analyses restricted to youth aged over 12 years showed similar patterns, with ADHD also differing significantly among adoptees (*p* = 0.017). Post-traumatic stress disorder (PTSD) showed a non-significant trend towards higher rates in FC (*p* = 0.101) (see Supplementary Table S1).

At discharge, most patients were prescribed psychotropic medication, most commonly antipsychotics (approximately 80–90% across groups), with no significant between-group differences in rates of prescription. In contrast, ADHD medication (*p* = 0.040) and mood stabilisers (*p* = 0.011) were prescribed more frequently among adopted youth (Table 3C).

### 3. Mental health service use before and after hospitalisation

Mental health service use before and after hospitalisation is summarised in Table 4. Length of inpatient stay did not differ significantly between groups (mean range: 16–19 days; *p* = 0.537).

**Table 4.** Mental health service use before and after hospitalisation.

| **Outcome** | **FC (n = 96)** | **Adoption (n = 60)** | **CWS (n = 80)** | **Test statistic** | **p-value** |
| --- | --- | --- | --- | --- | --- |
| **Length of hospital stay, days (mean ± SD [range])** | 16.6 ± 12.2 [3–82] | 16.9 ± 9.9 [2–61] | 18.9 ± 15.6 [2–86] | H = 1.24 | 0.537 |
| **Psychiatric readmission during follow-up, n (%)** | 46 (47.9%) | 38 (63.3%) | 36 (45.0%) | $\chi^2(2) = 5.17$ | 0.076 |
| **Emergency department visit after discharge, n (%)** | 71 (74.0%) | 53 (88.3%) | 52 (65.0%) | $\chi^2(2) = 9.88$ | 0.007 |
| **Time to first emergency department visit after discharge, days (median [95% CI])** | 44 [20.9–67.1] | 97 [50.1–143.9] | 117 [93.4–140.6] | Log-rank $\chi^2 = 1.48$ | 0.476 |
| **Time to psychiatric readmission, days (median [95% CI])** | 123 [97.9–148.1] | 132 [64.1–199.9] | 186 [78.8–293.2] | Log-rank $\chi^2 = 0.51$ | 0.776 |

Note. Values are n (%), mean ± SD [range], or median [95% CI], as appropriate. CI = confidence interval; $\chi^2$ = chi-square statistic; H = Kruskal–Wallis H statistic. Emergency department visit after discharge was defined as the occurrence of at least one emergency department visit following discharge. Between-group comparisons used chi-square tests for categorical variables, Kruskal–Wallis tests for continuous variables, and Kaplan–Meier survival analysis with log-rank tests for time-to-event outcomes.

The emergency department was the most common referral source overall and was particularly frequent among FC youth (57.9%), whereas outpatient services were the primary referral source for adopted youth (33.3%) and child welfare supervision youth (41.5%) (*p* < 0.001). Discharge destinations did not differ significantly between groups, with most youth referred to outpatient care (50.8–62.8%) or day hospital programmes (21.3–36.7%). Absconding during admission occurred exclusively among FC youth (5.2%; *p* = 0.024).

Psychiatric readmission during follow-up was most frequent among adopted youth (63.3%), compared with FC (47.9%) and youth under child welfare supervision (45.0%). Although the overall between-group comparison did not reach statistical significance (χ²(2) = 5.17, *p* = 0.076), logistic regression analyses indicated that adopted youth had significantly higher odds of readmission than youth in FC (β = 0.77*, p* = 0.022), while this did not differ significantly for youth under child welfare supervision (β = −0.27, *p* = 0.384). Median time to psychiatric readmission also did not differ significantly across groups (log-rank χ² = 0.51, *p* = 0.776; medians: FC 123 days, adoption 132 days, child welfare supervision 186 days; Figure 1).

**Figure 1.** Time to psychiatric readmission by group. Kaplan–Meier curves showing time to psychiatric readmission following discharge among youth in foster care, adoption, and child welfare supervision. Between-group differences were not statistically significant (log-rank χ² = 0.51, p = 0.776).

By contrast, emergency department re-consultation after discharge differed significantly between groups (χ²(2) = 9.88, *p* = 0.007), occurring most frequently among adopted youth (88.3%), followed by FC youth (74.0%), and least frequently among youth under child welfare supervision (65.0%). Kaplan–Meier survival analyses showed no significant between-group differences in time to first emergency department visit following discharge (log-rank χ² = 1.48, *p* = 0.476), although median time to first emergency department visit was shortest among FC youth (44 days; 95% CI 20.9–67.1), compared with adopted youth (97 days; 95% CI 50.1–143.9) and child welfare supervision youth (117 days; 95% CI 93.4–140.6) (Figure 2). Multilevel mixed-effects models indicated that group membership did not independently predict emergency department utilisation. However, emergency department use increased significantly after discharge (β = 2.67, *p* < 0.001), while higher emergency department utilisation prior to admission was associated with fewer subsequent emergency department visits (β = −0.85, *p* = 0.004).

**Figure 2.** Time to first emergency department contact after discharge by group. Kaplan–Meier curves showing time to first emergency department contact following discharge among youth in foster care, adoption, and child welfare supervision. Between-group differences were not statistically significant (log-rank χ² = 1.48, *p* = 0.476).

### 4. Clinical complexity

Clinical complexity was high across all groups (Table 5). In the full sample, the mean number of psychiatric diagnoses at discharge was 2.94 (SD = 1.68) among FC youth, 2.95 (SD = 1.56) among adopted youth, and 2.39 (SD = 1.36) among child welfare supervision youth. Between-group differences in the number of diagnoses approached statistical significance (Kruskal–Wallis χ² = 6.00, *p* = 0.050). Analyses restricted to youth aged over 12 years revealed a statistically significant difference in comorbidity burden, with adopted youth and FC youth showing higher mean numbers of diagnoses than youth under child welfare supervision (Kruskal–Wallis χ² = 6.15, *p* = 0.046).

**Table 5.**
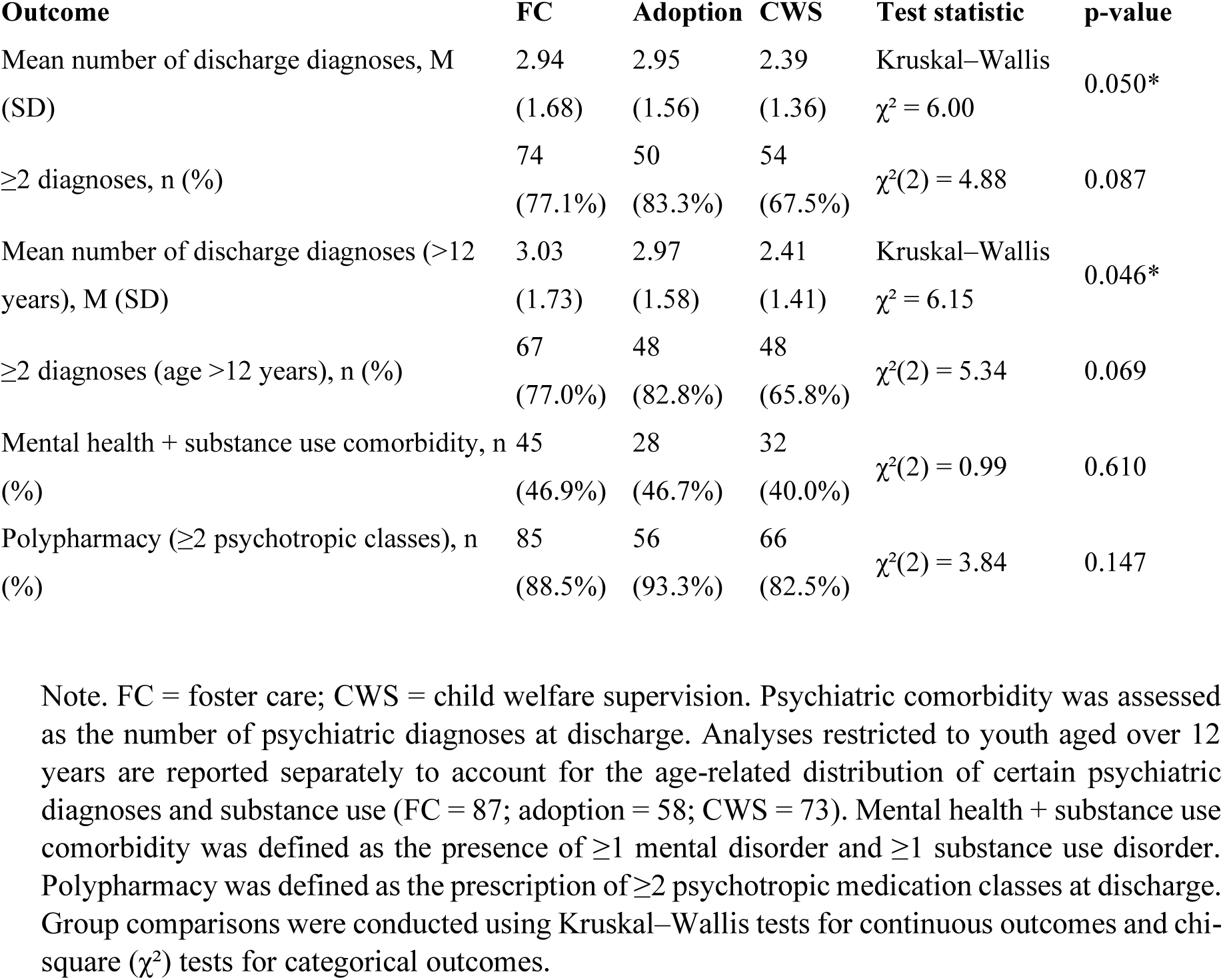
Clinical complexity and comorbidity across groups.

| **Table 5. Clinical complexity and comorbidity across groups** |  |  |  |  |  |
| --- | --- | --- | --- | --- | --- |
| **Outcome** | **FC** | **Adoption** | **CWS** | **Test statistic** | **p-value** |
| Mean number of discharge diagnoses, M (SD) | 2.94<br>(1.68) | 2.95<br>(1.56) | 2.39<br>(1.36) | Kruskal–Wallis<br>$\chi^2 = 6.00$ | 0.050* |
| $\geq 2$ diagnoses, n (%) | 74<br>(77.1%) | 50<br>(83.3%) | 54<br>(67.5%) | $\chi^2(2) = 4.88$ | 0.087 |
| Mean number of discharge diagnoses (>12 years), M (SD) | 3.03<br>(1.73) | 2.97<br>(1.58) | 2.41<br>(1.41) | Kruskal–Wallis<br>$\chi^2 = 6.15$ | 0.046* |
| $\geq 2$ diagnoses (age >12 years), n (%) | 67<br>(77.0%) | 48<br>(82.8%) | 48<br>(65.8%) | $\chi^2(2) = 5.34$ | 0.069 |
| Mental health + substance use comorbidity, n (%) | 45<br>(46.9%) | 28<br>(46.7%) | 32<br>(40.0%) | $\chi^2(2) = 0.99$ | 0.610 |
| Polypharmacy ($\geq 2$ psychotropic classes), n (%) | 85<br>(88.5%) | 56<br>(93.3%) | 66<br>(82.5%) | $\chi^2(2) = 3.84$ | 0.147 |

Note. FC = foster care; CWS = child welfare supervision. Psychiatric comorbidity was assessed as the number of psychiatric diagnoses at discharge. Analyses restricted to youth aged over 12 years are reported separately to account for the age-related distribution of certain psychiatric diagnoses and substance use (FC = 87; adoption = 58; CWS = 73). Mental health + substance use comorbidity was defined as the presence of $\geq 1$ mental disorder and $\geq 1$ substance use disorder. Polypharmacy was defined as the prescription of $\geq 2$ psychotropic medication classes at discharge. Group comparisons were conducted using Kruskal–Wallis tests for continuous outcomes and chi-square ( $\chi^2$ ) tests for categorical outcomes.

Overall, between 67.5% and 83.3% of youth presented with two or more psychiatric diagnoses. When comorbidity was dichotomised (≥2 vs. <2 diagnoses), between-group differences did not reach statistical significance in the full sample (χ²(2) = 4.88, *p* = 0.087) or in analyses restricted to youth aged over 12 years (χ²(2) = 5.34, *p* = 0.069).

Combined mental health and substance use comorbidity was also common, affecting 46.9% of FC youth, 46.7% of adopted youth, and 40.0% of child welfare supervision youth, with no significant between-group differences (χ²(2) = 0.99, *p* = 0.610).

Polypharmacy was widespread across the sample, with 87.7% of youth prescribed two or more psychotropic medication classes at discharge. Rates were highest among adopted youth (93.3%), followed by FC youth (88.5%) and youth under child welfare supervision (82.5%), although these differences were not statistically significant (χ²(2) = 3.84, *p* = 0.147).

Taken together, psychiatric comorbidity and polypharmacy were pervasive across all child protection groups. Adopted youth and FC youth showed a higher descriptive burden of clinical complexity than child welfare supervision youth, particularly among adolescents aged over 12 years; however, most between-group differences were modest and should be interpreted cautiously.

#### Supplementary Results

Further exploratory analyses examining associations between substance use and psychiatric diagnoses, length of stay by reason for admission, and associations between maltreatment history and psychiatric diagnoses across placement groups are presented in the Supplementary Material (Supplementary Tables S2–S3 and Supplementary Figures S1–S2).

## Discussion

To our knowledge, this is the first study to directly compare clinical complexity and mental health service use among psychiatrically hospitalised youth across the three main child protection arrangements: FC, adoption, and child welfare supervision. Building on prior research focused primarily on youth in FC (Solerdelcoll et al., 2022), this study broadens the scope to include adoptees and child welfare supervision youth, providing a more comprehensive framework for understanding how placement context and developmental history shape clinical profiles and service trajectories (Turney & Wildeman, 2017). Although all groups showed high levels of psychopathology, comorbidity, and service involvement, distinct and clinically meaningful patterns emerged, consistent with their differing exposure to adversity, developmental vulnerabilities, and placement pathways.

Youth in FC exhibited the highest prevalence of externalising and substance use disorders, alongside greater trauma exposure and more frequent presentations with behavioural dysregulation at admission. The FC group also included a proportion of unaccompanied migrant minors, a population frequently exposed to multiple adversities before and during migration that may contribute to increased mental health vulnerability. These findings are consistent with prior evidence indicating that cumulative adversity, disrupted attachment relationships, and placement instability contribute to emotional and behavioural dysregulation (Ford et al., 2007; Kerr-Davis et al., 2023; Lane et al., 2023; Lehmann, 2021; Maguire et al., 2024; McWey et al., 2010; Meltzer et al., 2003). Notably, almost one-quarter of youth in FC entered care for the first time following psychiatric hospitalisation. This finding suggests that psychiatric crises may reveal previously unrecognised situations of vulnerability and prompt child protection intervention, underscoring the close interconnection between mental health needs and child protection involvement. Their acute behavioural presentations were also reflected in shorter inpatient admissions and a higher likelihood of admission via the emergency department, underscoring the crisis-driven nature of service use in this group. Although overall service use and length of hospitalisation did not differ significantly between groups, FC youth had the shortest observed median time to emergency department re-consultation after discharge. This descriptive pattern may suggest challenges in sustained clinical stabilisation or continuity of care (Kelly et al., 2026; Soleimani et al., 2016).

In contrast, adopted adolescents showed a higher prevalence of neurodevelopmental and prenatal adversity-related conditions, including ASD, FASD, and prematurity. These patterns are consistent with evidence highlighting the enduring effects of early biological vulnerability and, for some, early institutional deprivation, even when subsequent caregiving environments are stable (Palacios & Brodzinsky, 2010; Rodriguez-Perez et al., 2023; Sonuga-Barke et al., 2017). Clinically, adopted youth tended to present with fewer acute behavioural crises but greater developmental and regulatory complexity, often involving cognitive, emotional, and attachment-related difficulties. The higher odds of psychiatric readmission observed in this group suggest persistent challenges in maintaining outpatient stability, despite generally stable and consistent family involvement. Although between-group differences in psychotropic polypharmacy were not statistically significant, the high rates of psychotropic polypharmacy among adopted youth further underscore the treatment complexity characteristic of this population.

Youth under child welfare supervision exhibited comparatively lower diagnostic complexity but a predominance of internalising symptoms, particularly affective and suicidal presentations. Remaining within the biological families under protective supervision did not preclude clinically significant psychopathology, suggesting that persistent psychosocial stressors and delayed access to specialised intervention may contribute to sustained internalising distress (Burns et al., 2004; Jud et al., 2016; Lane et al., 2023). Although readmission rates were lower in this group, the prominence of affective and self-harm-related presentations raises concern that internalising difficulties may be under-recognised or insufficiently addressed within community-based services, particularly in the absence of overt behavioural disruption.

Clinical complexity—operationalised through diagnostic comorbidity and psychotropic polypharmacy—was pervasive across all CPS groups, reflecting the substantial severity of psychopathology among youth requiring inpatient care. Contrary to our initial hypotheses, adopted adolescents showed the highest rates of psychiatric comorbidity and pharmacological complexity, although these differences did not reach statistical significance. This pattern suggests that early neurodevelopmental and prenatal vulnerabilities may contribute to clinical complexity to a degree comparable with the cumulative psychosocial adversity associated with foster care placements (dosReis et al., 2011; Keyes et al., 2008; Ligier et al., 2022). Taken together, these findings challenge the assumption that residential care exposure alone accounts for elevated treatment burden and emphasise the importance of developmentally informed, multidisciplinary treatment models across CPS populations (Cawthorne & Woolgar, 2025; Radel et al., 2023).

Exploratory analyses further highlighted the context-dependent impact of maltreatment. Among FC youth, trauma exposure was associated with PTSD and suicidality, consistent with the cumulative effects of chronic adversity and placement disruption (Greeson et al., 2011; Haselgruber et al., 2021). Among adoptees, maltreatment—although less frequently documented—was associated with particularly complex outcomes, including PTSD, conduct disorder, and FASD, suggesting enduring neurodevelopmental and behavioural sequelae of early trauma despite subsequent placement stability (Brodzinsky et al., 2022; Solerdelcoll et al., 2026). In contrast, associations between maltreatment and diagnosis were less evident among youth under child welfare supervision, potentially reflecting underreporting, differences in case trajectories, or variability in the timing and chronicity of adversity. Together, these findings suggest that the developmental consequences of maltreatment may be shaped not only by exposure risk but also by the timing, context, and stability of subsequent caregiving environments (Berens et al., 2017; Gilbert et al., 2009).

Overall, child protection status may interact with developmental history, placement stability, and systemic support in shaping patterns of psychiatric presentation and service use. Youth in FC appeared particularly vulnerable to crisis-driven behavioural dysregulation and showed a pattern of earlier post-discharge emergency re-consultation; adopted youth exhibited complex neurodevelopmental profiles and higher odds of psychiatric readmission; and youth under child welfare supervision showed predominantly internalising presentations that may be less readily recognised in family-based contexts.

These differentiated patterns highlight the importance of placement-sensitive mental health strategies that bridge inpatient and community care and address the specific vulnerabilities associated with each child protection pathway (Solerdelcoll et al., 2026).

## Clinical implications

This study underscores the need for differentiated, trauma-informed, and developmentally attuned approaches to inpatient and post-discharge care. FC youth may benefit from targeted interventions focusing on behavioural regulation and substance use prevention, combined with structured discharge planning to reduce crisis recidivism. Adopted adolescents may require neurodevelopmentally informed, family-based treatment plans that integrate attachment-focused strategies and enhanced parental support to address regulatory and relational difficulties. For youth under child welfare supervision, systematic and proactive screening for affective symptoms and suicidality is important, as difficulties may be underestimated in family-based contexts. Across all groups, sustained coordination between mental health and child protection services remains essential to ensure continuity of care, minimise fragmentation, and promote greater stability following hospitalisation (McKenzie et al., 2025).

## Strengths and Limitations

To our knowledge, this study is among the first to directly compare hospitalised youth admitted to a psychiatric unit across the three main child protection pathways, providing a broader framework for understanding how placement context shapes mental health needs and service trajectories. Strengths include the use of detailed, real-world clinical data from a well-characterised inpatient cohort, together with the systematic integration of diagnostic, service-use, and contextual variables.

Several limitations should be acknowledged. First, the retrospective design meant that key psychosocial variables (e.g. early adversity, prenatal exposures, family psychiatric history) were often unavailable or inconsistently documented, particularly among adopted youth, limiting the interpretation of between-group comparisons for these variables. Second, maltreatment exposure was derived from clinical records rather than assessed using standardised instruments, which may have resulted in underreporting or variability in documentation. Psychiatric diagnoses were established by consultant child and adolescent psychiatrists during routine inpatient care, although some variability in diagnostic ascertainment cannot be excluded.

As in previous work (Solerdelcoll et al., 2022), this study focused on youth admitted to a psychiatric unit. Therefore, the findings should be interpreted within the context of severe mental illness and may not be generalisable to child protection populations receiving community-based care. Finally, this single-site study was conducted in a public tertiary hospital in Catalonia, and the findings may not be fully generalisable to other healthcare or CPS. The study period also included the COVID-19 pandemic and associated lockdown measures, which may have influenced patterns of psychiatric presentation and service utilisation. However, all groups were recruited from the same inpatient unit throughout the study period, reducing the likelihood that pandemic-related changes systematically biased between-group comparisons. Future multi-site prospective studies using standardised, multi-informant measures and longitudinal follow-up are needed to confirm and extend these findings.

## Conclusions

This study provides the first direct comparison of psychiatric presentations and service trajectories among psychiatrically hospitalised youth across FC, adoption, and child welfare supervision. Although all groups demonstrated severe and complex psychopathology, their clinical profiles diverged in clinically meaningful ways, shaped by developmental history, placement context, and continuity of care. Youth in FC more frequently presented with behavioural dysregulation, substance use, and trauma exposure, and showed a pattern of earlier post-discharge emergency department contact that may reflect heightened vulnerability to recurrent crises. Adopted youth exhibited higher rates of neurodevelopmental and prenatal adversity-related conditions and showed higher odds of psychiatric readmission, despite living in stable family environments. Youth under child welfare supervision, though less diagnostically complex overall, showed prominent affective and suicidal symptoms that may be under-recognised in family-based settings.

These findings indicate that placement context does not uniformly mitigate psychiatric vulnerability among CPS-involved youth and underscore the importance of trauma-informed, placement-sensitive, and developmentally tailored approaches to inpatient and post-discharge care. Accordingly, care models should strengthen continuity between inpatient and community services while addressing the distinct needs associated with each child protection pathway.

Future research should examine long-term clinical and functional outcomes and evaluate interventions designed to promote sustained recovery and stability across child protection pathways.

## Supporting information

Supplementary Results, Tables S1-S3 and Figures S1-S2

## Acknowledgements

The authors thank all staff of the Department of Child and Adolescent Psychiatry and Psychology at Hospital Clínic de Barcelona for their support and commitment to patient care. The authors also thank 3Datos for statistical assistance.

## Author contributions

MS conceptualised and designed the study, collected the data, performed the statistical analyses, interpreted the findings, drafted the manuscript and revised it. GS and IB conceptualised and designed the study, supervised the project, contributed to the interpretation of the findings and critically revised the manuscript for important intellectual content. All authors approved the final version of the manuscript and agree to be accountable for all aspects of the work.

## Ethics approval and consent to participate

The study was approved by the Clinical Research Ethics Committee (CEIm) of Hospital Clínic de Barcelona (HCB/2018/0111) and was conducted in accordance with the Declaration of Helsinki and applicable local regulations. Given the retrospective design of the study, the requirement for informed consent was waived by the CEIm.

## Consent for publication

Not applicable.

## Data availability

The datasets generated and/or analysed during the current study are not publicly available because they contain sensitive clinical information. They may be available from the corresponding author on reasonable request, subject to institutional approval and applicable ethical and data-protection regulations.

## Code availability

Not applicable.

## Declaration of generative AI use

During the preparation of this work, the authors used ChatGPT (OpenAI) to assist with language editing and improving readability. Following use of this tool, the authors reviewed and edited the content as necessary and take full responsibility for the content of the manuscript.

## Abbreviations

CPS: child protection system
FC: foster care
ASD: autism spectrum disorder
FASD: foetal alcohol spectrum disorder
DSM-5: Diagnostic and Statistical Manual of Mental Disorders, Fifth Edition
IQR: interquartile range
ANOVA: analysis of variance
CI: confidence interval
ADHD: attention-deficit/hyperactivity disorder
PTSD: post-traumatic stress disorder.

