## Supplementary Results, Tables S1-S3 and Figures S1-S2 for "Psychiatric Characteristics and Mental Health Service Use Among Hospitalised Youth in Foster Care, Adoption, and Child Welfare Supervision: A Retrospective Comparative Study"

**S1. Associations Between Psychiatric Diagnoses and Substance Use**

*(See Supplementary Table S2)*

Substance use (tobacco, cannabis, alcohol, and other substances) showed consistent associations with affective and behavioural disorders across the sample. Tobacco use was associated with ADHD ($p$ = .035), psychotic disorders ($p$ = .023), mood disorders ($p$ = .001), and conduct disorder ($p$ < .001). Cannabis use was associated with mood disorders ($p$ = .001) and conduct disorder ($p$ < .001). Alcohol consumption was associated with foetal alcohol spectrum disorder (FASD) (p = .002), mood disorders ($p$ = .002), and conduct disorder ($p$ = .002). Use of other substances was associated with conduct disorder ($p$ = .003). Across all substance categories, ASD was significantly less frequent among substance users than non-users (all $p$ < .05).

Exploratory analyses showed broadly similar diagnostic–substance use associations across placement groups (FC, adoption, and CWS). The only significant exception concerned ADHD, for which other substance use was positively associated with the disorder among youth in CWS ($p$ = .032), contrasting with the overall pattern of lower ADHD prevalence among substance users.

**S2. Length of Stay by Primary Reason for Admission**

*(See Supplementary Figure S1)*

Length of hospitalisation varied significantly by primary reason for admission (F = 3.43, $p$ = .002, $\eta^2$ = .095). Eating disorders were associated with the longest admissions (M = 39.5 ± 34.2 days), followed by mood disorders (22.0 ± 18.5 days) and psychotic symptoms (21.3 ± 17.0 days). Intermediate lengths of stay were observed for suicidal ideation or attempt (18.1 ± 12.9 days) and heteroaggressive behaviour (17.7 ± 12.5 days). Behavioural dysregulation, the most frequent reason for admission, was associated with the shortest stays (14.4 ± 7.1 days). No significant differences were observed for secondary admission reasons ($p$ = .799).

**S3. Maltreatment and Psychiatric Diagnoses by Placement Group**

*(See Supplementary Table S3 and Supplementary Figure S2)*

The association between documented maltreatment history and psychiatric diagnoses was examined within each placement group.

Among youth in FC, maltreatment was associated with higher rates of PTSD (34.9% vs. 6.3%, p = .024, Cramer's V = .254) and with admission for suicidal ideation or attempts (31.7% vs. 6.3%). Conduct and mood disorders did not reach statistical significance but showed small effect sizes.

Among adopted youth, maltreatment—although less frequently documented—was associated with higher rates of PTSD (41.2% vs. 4.2%, *p* = .003, V = .460), conduct disorder (41.2% vs. 12.5%, *p* = .035, V = .329), and FASD (35.3% vs. 4.2%, *p* = .009, V = .408).

In the CWS group, ADHD (33.3% vs. 5.5%, p = .002), oppositional defiant disorder (27.8% vs. 7.3%, p = .022) and intellectual disability (38.9% vs. 7.3%, p = .001) were significantly more prevalent among youth without documented maltreatment. Associations involving ASD were non-significant within all three placement groups.

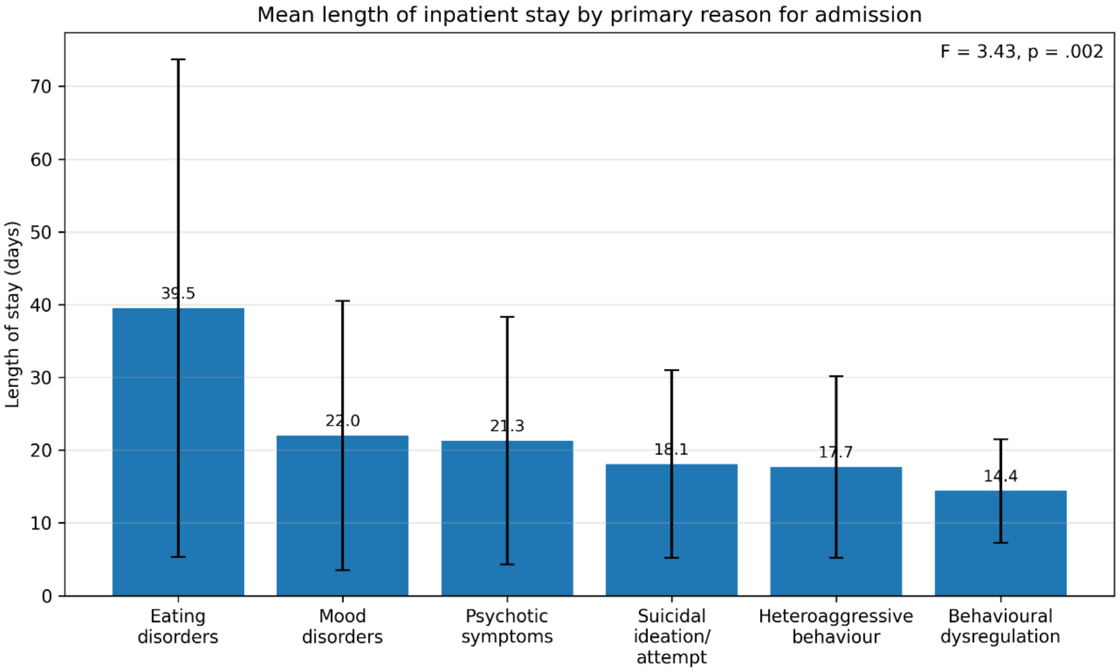


**Figure S1.** Mean length of inpatient stay by primary reason for admission among psychiatrically hospitalised youth. Error bars represent ±1 standard deviation. Length of stay differed significantly across admission categories (F = 3.43, *p* = .002, $\eta^2 = .095$).

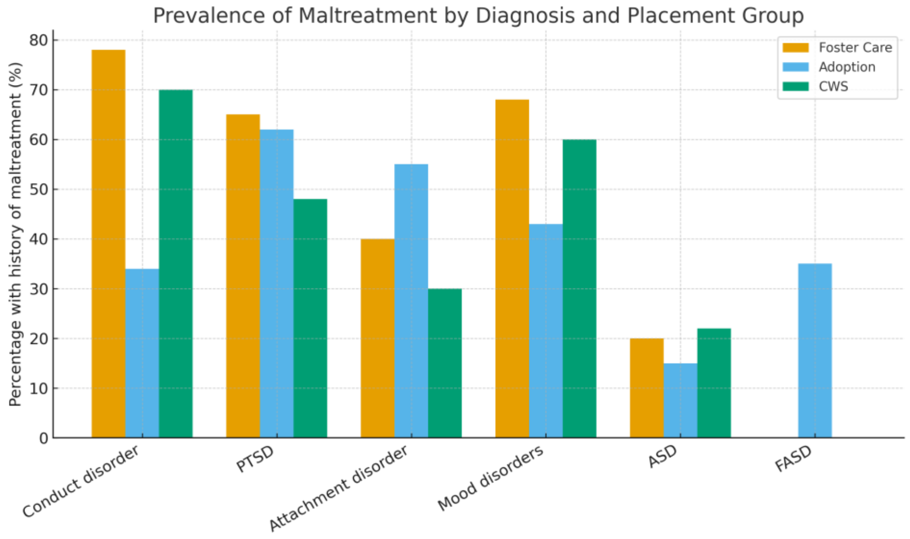


**Figure S2.** Prevalence of maltreatment history among psychiatrically hospitalised youth by diagnosis and placement group.

**Supplementary Table S1. Psychiatric diagnoses and substance use in the full sample and among youth aged >12 years**

| Diagnosis | FC (All) | Adoptees (All) | CWS (All) | $\chi^2$ | *p* | V | FC (>12 years) | Adoptees (>12 years) | CWS (>12 years) | $\chi^2$ | *p* | V |
|---|---|---|---|---|---|---|---|---|---|---|---|---|
| ADHD | 17 (17.7%) | 17 (28.3%) | 11 (13.8%) | 4.92 | 0.085 | 0.14 | 13 (14.9%) | 17 (29.3%) | 8 (11.0%) | 8.19 | 0.017 | 0.19 |
| Any psychotic disorder | 12 (12.5%) | 7 (11.7%) | 12 (15.0%) | 0.39 | 0.822 | 0.04 | 11 (12.6%) | 7 (12.1%) | 11 (15.1%) | 0.31 | 0.858 | 0.04 |
| Clinical high-risk state for psychosis | 3 (3.1%) | 3 (5.0%) | 1 (1.3%) | 1.69 | 0.430 | 0.08 | 3 (3.4%) | 3 (5.2%) | 1 (1.4%) | 1.53 | 0.465 | 0.08 |
| Any affective disorder | 38 (39.6%) | 24 (40.0%) | 37 (46.3%) | 0.92 | 0.631 | 0.06 | 35 (40.2%) | 24 (41.4%) | 35 (47.9%) | 1.06 | 0.588 | 0.07 |
| ODD | 7 (7.3%) | 4 (6.7%) | 9 (11.3%) | 1.22 | 0.543 | 0.07 | 7 (8.0%) | 4 (6.9%) | 9 (12.3%) | 1.37 | 0.505 | 0.08 |
| **ASD** | **13 (13.5%)** | **16 (26.7%)** | **6 (7.5%)** | **10.18** | **0.006** | **0.21** | **10 (11.5%)** | **14 (24.1%)** | **5 (6.8%)** | **8.79** | **0.012** | **0.20** |
| Intellectual disability | 10 (10.4%) | 11 (18.3%) | 13 (16.3%) | 2.21 | 0.331 | 0.10 | 8 (9.2%) | 9 (15.5%) | 10 (13.7%) | 1.46 | 0.483 | 0.08 |
| **Conduct disorder** | **30 (31.2%)** | **17 (28.3%)** | **12 (15.0%)** | **6.62** | **0.036** | **0.17** | **28 (32.2%)** | **17 (29.3%)** | **9 (12.3%)** | **9.27** | **0.010** | **0.21** |
| Eating disorder | 3 (3.1%) | 4 (6.7%) | 4 (5.0%) | 1.07 | 0.585 | 0.07 | 3 (3.4%) | 4 (6.9%) | 4 (5.5%) | 0.91 | 0.635 | 0.06 |
| OCD | 0 (0.0%) | 1 (1.7%) | 1 (1.2%) | 1.45 | 0.483 | 0.08 | 0 (0.0%) | 1 (1.7%) | 0 (0.0%) | 2.77 | 0.250 | 0.11 |
| PTSD | 23 (24.0%) | 8 (13.3%) | 11 (13.8%) | 4.20 | 0.122 | 0.13 | 21 (24.1%) | 7 (12.1%) | 10 (13.7%) | 4.58 | 0.101 | 0.15 |
| Tourette syndrome | 1 (1.0%) | 2 (3.3%) | 0 (0.0%) | 3.10 | 0.212 | 0.12 | 1 (1.1%) | 2 (3.4%) | 0 (0.0%) | 2.89 | 0.236 | 0.12 |
| Personality disorder | 4 (4.2%) | 4 (6.7%) | 7 (8.8%) | 1.55 | 0.460 | 0.08 | 4 (4.6%) | 4 (6.9%) | 7 (9.6%) | 1.54 | 0.462 | 0.08 |
| Tobacco use | 38 (39.6%) | 22 (36.7%) | 28 (35.0%) | 0.41 | 0.817 | 0.04 | 37 (42.5%) | 22 (37.9%) | 28 (38.4%) | 0.42 | 0.812 | 0.04 |
| Cannabis use | 37 (38.5%) | 21 (35.0%) | 25 (31.2%) | 1.02 | 0.601 | 0.07 | 37 (42.5%) | 21 (36.2%) | 25 (34.2%) | 1.27 | 0.530 | 0.08 |
| **Alcohol use** | **21 (21.9%)** | **13 (21.7%)** | **7 (8.8%)** | **6.27** | **0.044** | **0.16** | **21 (24.1%)** | **13 (22.4%)** | **7 (9.6%)** | **6.18** | **0.046** | **0.17** |
| **Other substance use** | **25 (26.0%)** | **3 (5.0%)** | **7 (8.8%)** | **16.49** | **< 0.001** | **0.26** | **25 (28.7%)** | **3 (5.2%)** | **7 (9.6%)** | **17.74** | **< 0.001** | **0.29** |

Note. Diagnoses are not mutually exclusive. Values are *n* (%). $\chi^2$ = chi-square statistic. Effect sizes are reported as Cramer's V.

FC = foster care; CWS = child welfare supervision; ADHD = attention-deficit/hyperactivity disorder; ODD = oppositional defiant disorder; ASD = autism spectrum disorder; OCD = obsessive-compulsive disorder; PTSD = post-traumatic stress disorder.

**Supplementary Table S2. Associations between substance use and psychiatric discharge diagnoses among psychiatrically hospitalised youth.**

| Discharge diagnosis | Tobacco use | | | | | Cannabis use | | | | | Alcohol use | | | | | Other substance use | | | | |
|---|---|---|---|---|---|---|---|---|---|---|---|---|---|---|---|---|---|---|---|---|
| | No *(n=147)* | Yes *(n=88)* | $\chi^2$ | *p* | *V* | No *(n=153)* | Yes *(n=83)* | $\chi^2$ | *p* | *V* | No *(n=195)* | Yes *(n=41)* | $\chi^2$ | *p* | *V* | No *(n=200)* | Yes *(n=35)* | $\chi^2$ | *p* | *V* |
| ADHD | 22 (15.0%) | 23 (26.1%) | 4.44 | .035* | .137(p) | 25 (16.3%) | 20 (24.1%) | 2.10 | .148 | .094(t) | 37 (19.0%) | 8 (19.5%) | .01 | .936 | .005(t) | 41 (20.5%) | 4 (11.4%) | 1.58 | .208 | .082(t) |
| Any psychotic condition | 17 (11.6%) | 20 (22.7%) | 5.17 | .023* | .148(p) | 21 (13.7%) | 17 (20.5%) | 1.82 | .178 | .088(t) | 30 (15.4%) | 8 (19.5%) | .43 | .513 | .043(t) | 28 (14.0%) | 10 (28.6%) | 4.67 | .031* | .141(p) |
| Mood disorders | | | 21.10 | .001* | .300(m) | | | 19.80 | .001* | .29(p) | | | 18.42 | .002* | .279(p) | | | 11.68 | .039* | .223(p) |
| Adjustment disorder | 20 (13.6%) | 8 (9.1%) | | | | 22 (14.4%) | 6 (7.2%) | | | | 27 (13.8%) | 1 (2.4%) | | | | 24 (12%) | 3 (8.6%) | | | |
| Depressive disorder | 31 (21.1%) | 4 (4.5%) | | | | 30 (19.6%) | 5 (6.0%) | | | | 34 (17.4%) | 1 (2.4%) | | | | 34 (17%) | 1 (2.9%) | | | |
| Bipolar disorder | 3 (2.0%) | 4 (4.5%) | | | | 3 (2.0%) | 4 (4.8%) | | | | 3 (1.5%) | 4 (9.8%) | | | | 6 (3.0%) | 1 (2.9%) | | | |
| MED | 16 (10.9%) | 4 (4.5%) | | | | 15 (9.8%) | 5 (6.0%) | | | | 15 (7.7%) | 5 (12.2%) | | | | 19 (9.5%) | 1 (2.9%) | | | |
| DMDD | 6 (4.1%) | 3 (3.4%) | | | | 8 (5.2%) | 1 (1.2%) | | | | 7 (3.6%) | 2 (4.9%) | | | | 9 (4.5%) | - | | | |
| Any mood disorder | 76 (51.7%) | 23 (26.1%) | 14.76 | <.001* | .251(p) | 78 (51.0%) | 21 (25.3%) | 14.57 | <.001* | .248(p) | 86 (44.1%) | 13 (31.7%) | 2.14 | .144 | .095(t) | 92 (46.0%) | 6 (17.1%) | 10.20 | .001* | .208(p) |
| ODD | 9 (6.1%) | 11 (12.5%) | 2.87 | .090 | .111(p) | 9 (5.9%) | 11 (13.3%) | 3.77 | .052 | .126(p) | 17 (8.7%) | 3 (7.3%) | .09 | .770 | .019(t) | 19 (9.5%) | 1 (2.9%) | 1.69 | .194 | .085(t) |
| ASD | 30 (20.4%) | 2 (2.3%) | 15.39 | <.001* | .256(p) | 28 (18.3%) | 4 (4.8%) | 8.34 | .004* | .188(p) | 31 (15.9%) | 1 (2.4%) | 5.23 | .022* | .149(p) | 31 (15.5%) | 1 (2.9%) | 4.05 | .044* | .131(p) |
| Intellectual disability | 23 (15.6%) | 11 (12.5%) | .44 | .507 | .043(t) | 26 (17.0%) | 8 (9.6%) | 2.36 | .124 | .100(p) | 31 (15.9%) | 3 (7.3%) | 2.02 | .155 | .093(t) | 31 (15.5%) | 3 (8.6%) | 1.16 | .282 | .07(t) |
| Conduct disorder | 21 (14.3%) | 34 (38.6%) | 18.21 | <.001* | .278(p) | 22 (14.4%) | 33 (39.8%) | 19.39 | <.001* | .287(p) | 38 (19.5%) | 17 (41.5%) | 9.15 | .002* | .197(p) | 40 (20.0%) | 15 (42.9%) | 8.68 | .003* | .192(p) |
| Eating disorder | 9 (6.1%) | 2 (2.3%) | 1.83 | .176 | .088(t) | 9 (5.9%) | 2 (2.4%) | 1.46 | .227 | .079(t) | 9 (4.6%) | 2 (4.9%) | .01 | .942 | .005(t) | 10 (5.0%) | 1 (2.9%) | .31 | .580 | .036(t) |
| OCD | 2 (1.4%) | - | 1.21 | .272 | .072(t) | 2 (1.3%) | - | 1.09 | .296 | .068(t) | 2 (1.0%) | - | .42 | .515 | .042(t) | 2 (1.0%) | - | .35 | .552 | .039(t) |
| PTSD | 29 (19.7%) | 13 (14.8%) | .92 | .337 | .063(t) | 29 (19.0%) | 13 (15.7%) | .40 | .528 | .041(t) | 33 (16.9%) | 9 (22%) | .58 | .444 | .05(t) | 38 (19.0%) | 4 (11.4%) | 1.16 | .281 | .07(t) |
| Tourette Syndrome | 3 (2%) | - | 1.82 | .177 | .088(t) | 2 (1.3%) | 1 (1.2%) | .01 | .947 | .004(t) | 3 (1.5%) | - | .64 | .424 | .052(t) | 3 (1.5%) | - | .53 | .466 | .048(t) |
| Attachment disorder | 8 (5.4%) | 2 (2.3%) | 1.36 | .244 | .076(t) | 7 (4.6%) | 3 (3.6%) | .12 | .726 | .023(t) | 8 (4.1%) | 2 (4.9%) | .05 | .823 | .015(t) | 10 (5.0%) | - | 1.83 | .176 | .088(t) |
| FASD | 5 (3.4%) | 7 (8.0%) | 2.35 | .125 | .101(p) | 6 (3.9%) | 6 (7.2%) | 1.22 | .269 | .072(t) | 6 (3.1%) | 6 (14.6%) | 9.38 | .002* | .199(p) | 12 (6.0%) | - | 2.21 | .137 | .097(t) |
| Medical/perinatal conditions | 24 (16.3%) | 8 (9.1%) | 2.45 | .118 | .102(p) | 25 (16.3%) | 7 (8.4%) | 2.87 | .090 | .110(p) | 28 (14.4%) | 4 (9.8%) | .61 | .434 | .051(t) | 29 (14.5%) | 3 (8.6%) | 0.89 | .345 | .062(t) |

**Note.** Any psychotic condition includes psychotic disorders and clinical high-risk state for psychosis. ADHD = attention-deficit/hyperactivity disorder; MED = mixed anxiety and depressive disorder; DMDD = disruptive mood dysregulation disorder; ODD = oppositional defiant disorder; ASD = autism spectrum disorder; OCD = obsessive–compulsive disorder; PTSD = post-traumatic stress disorder; FASD = foetal alcohol spectrum disorder. Medical/perinatal conditions include organic neurological disorders, malnutrition, prematurity, perinatal complications and other medical conditions. Low-frequency diagnoses were grouped into this variable to avoid sparse cells in $\chi^2$ analyses. $\chi^2$ = chi-square test; V = Cramer's V effect size (t = trivial, p = small, m = medium, g = large*)*.
$^*p < .05$.

**Supplementary Table S3. Primary reasons for admission and discharge diagnoses according to maltreatment status within placement groups (foster care, adoption, and child welfare supervision).**

| | Foster Care | | | | | Adoption | | | | | Child welfare supervision | | | | |
|---|---|---|---|---|---|---|---|---|---|---|---|---|---|---|---|
| | Maltreatment history | | | | | Maltreatment history | | | | | | Maltreatment history | | | |
| | No *(n=16)* | Yes *(n=63)* | $\chi^2$ | *p* | *V* | No *(n=24)* | Yes *(n=17)* | $\chi^2$ | *p* | *V* | No *(n=18)* | Yes *(n=55)* | $\chi^2$ | *p* | *V* |
| Primary reason for admission | | | 11.68 | .069 | .385(m) | | | .42 | .995 | .101(p) | | | 11.10 | .134 | .390(m) |
| Behavioural dysregulation | 9 (56.3%) | 24 (38.1%) | | | | 9 (37.5%) | 7 (41.2%) | | | | 7 (38.9%) | 11 (20.0%) | | | |
| Psychotic symptoms | 0 (0.0%) | 5 (7.9%) | | | | 2 (8.3%) | 1 (5.9%) | | | | 1 (5.6%) | 4 (7.3%) | | | |
| Suicidal ideation / attempt | 1 (6.3%) | 20 (31.7%) | | | | 7 (29.2%) | 4 (23.5%) | | | | 2 (11.1%) | 24 (43.6%) | | | |
| Heteroaggressive behaviour | 3 (18.8%) | 5 (7.9%) | | | | 2 (8.3%) | 2 (11.8%) | | | | 1 (5.6%) | 5 (9.1%) | | | |
| Affective symptoms | 2 (12.5%) | 6 (9.5%) | | | | 3 (12.5%) | 2 (11.8%) | | | | 3 (16.7%) | 5 (9.1%) | | | |
| Eating disorder | 0 (0.0%) | 0 (0.0%) | | | | 0 (0.0%) | 0 (0.0%) | | | | 1 (5.6%) | 2 (3.6%) | | | |
| Self-harm | 0 (0.0%) | 3 (4.8%) | | | | 0 (0.0%) | 0 (0.0%) | | | | 0 (0.0%) | 2 (3.6%) | | | |
| Substance use disorder | 1 (6.3%) | 0 (0.0%) | | | | 1 (4.2%) | 1 (5.9%) | | | | 3 (16.7%) | 2 (3.6%) | | | |
| Discharge diagnoses | | | | | | | | | | | | | | | |
| ADHD | 5 (31.3%) | 11 (17.5%) | 1.50 | .220 | .138(p) | 7 (29.2%) | 4 (23.5%) | .16 | .688 | .063(t) | 6 (33.3%) | 3 (5.5%) | 8.75 | .002* | .365(m) |
| Any psychotic disorder | 11 (68.8%) | 0 (0.0%) | Fisher | < .001* | - | 6 (25.0%) | 2 (11.8%) | 1.11 | .292 | .165(p) | 2 (11.1%) | 9 (16.4%) | .29 | .589 | .063(t) |
| Any mood disorder | 6 (37.5%) | 28 (44.4%) | .25 | .616 | .056(t) | 7 (29.2%) | 6 (35.3%) | .17 | .678 | .065(t) | 8 (44.4%) | 27 (49.1%) | .12 | .732 | .040(t) |
| ODD | 3 (18.8%) | 4 (6.3%) | 2.43 | .119 | - | 3 (12.5%) | 0 (0.0%) | 2.29 | .130 | .236(p) | 5 (27.8%) | 4 (7.3%) | 5.27 | .022* | .269(p) |
| ASD | 3 (18.8%) | 10 (15.9%) | .08 | .782 | .031(t) | 7 (29.2%) | 3 (17.6%) | .72 | .397 | .132(p) | 3 (16.7%) | 3 (5.5%) | 2.26 | .133 | .176(p) |
| Intellectual disability | 2 (12.5%) | 6 (9.5%) | .12 | .725 | .04(t) | 2 (8.3%) | 5 (29.4%) | 3.12 | .077 | .276(p) | 7 (38.9%) | 4 (7.3%) | 10.59 | .001* | .381(m) |
| Conduct disorder | 5 (31.3%) | 13 (20.6%) | .82 | .366 | .102(p) | 3 (12.5%) | 7 (41.2%) | 4.44 | .035* | .329(m) | 4 (22.2%) | 5 (9.1%) | 2.16 | .141 | .172(p) |
| Eating disorder | 0 (0.0%) | 2 (3.2%) | .52 | .470 | .081(t) | 2 (8.3%) | 1 (5.9%) | .09 | .767 | .046(t) | 1 (5.6%) | 3 (5.5%) | .01 | .987 | .002(t) |
| OCD | 0 (0.0%) | 0 (0.0%) | - | - | - | 0 (0.0%) | 0 (0.0%) | - | - | - | 1 (5.6%) | 0 (0.0%) | 3.10 | .078 | .206(p) |
| PTSD | 1 (6.3%) | 22 (34.9%) | 5.08 | .024* | .254(p) | 1 (4.2%) | 7 (41.2%) | 8.68 | .003* | .46(m) | 0 (0.0%) | 10 (18.2%) | 3.79 | .051 | .228(p) |
| Tourette Syndrome | 1 (6.3%) | 0 (0%) | 3.99 | .046* | .225(p) | 1 (4.2%) | 0 (0.0%) | .73 | .394 | .133(p) | 0 (0.0%) | 0 (0.0%) | - | - | - |
| Attachment disorder | 1 (6.3%) | 3 (4.8%) | .06 | .808 | .027(t) | 2 (8.3%) | 2 (11.8%) | .13 | .715 | .057(t) | 0 (0.0%) | 0 (0.0%) | - | - | - |
| FASD | 0 (0.0%) | 1 (1.6%) | .26 | .612 | .057(t) | 1 (4.2%) | 6 (35.3%) | 6.81 | .009* | .408(m) | 0 (0.0%) | 0 (0.0%) | - | - | - |
| Medical/perinatal conditions | 0 (0.0%) | 4 (6.3%) | Fisher | .577 | - | 9 (37.5%) | 8 (47.1%) | .37 | .540 | .096(t) | 2 (11.1%) | 7 (12.7%) | Fisher | 1.000 | - |

Note. ADHD = attention-deficit/hyperactivity disorder; ODD = oppositional defiant disorder; ASD = autism spectrum disorder; OCD = obsessive–compulsive disorder; PTSD = post-traumatic stress disorder; FASD = foetal alcohol spectrum disorder. Medical/perinatal conditions include organic neurological disorders, malnutrition, prematurity, perinatal complications, and other medical conditions.
Primary reason for admission was analysed using chi-square tests comparing category distributions within each placement group. Discharge diagnoses were analysed using chi-square tests or Fisher's exact test as appropriate. Cramer's V is reported as a measure of effect size (t = trivial, p = small, m = medium, g = large).
*$p < .05$.
